# An After-Action Review of COVID-19 Cases and Mitigation Measures at US Mission India, March 2020-July 2021

**DOI:** 10.1101/2023.08.24.23294557

**Authors:** Jaspreet Singh, Rajesh Yadav, Samantha Robinson, Mark Vanelli, Melissa Nyendak, Meghna Desai

## Abstract

**Introduction:** Between March 2020-June 2021, over 30 million COVID-19 cases were reported in India. We assessed the COVID-19 response across the US Mission India (US Embassy New Delhi, US Consulates – Mumbai, Hyderabad, Chennai and Kolkata) to plan future mitigation efforts and fill gaps in knowledge about COVID-19 transmission in a unique community like the US Mission.

**Method:** We described COVID-19 mitigation activities undertaken by the five US Mission India posts and conducted a secondary analysis of case investigation and contact tracing program data collected by the Health Unit from March 2020–July 2021.

**Results:** US Mission in India, in collaboration with multiple internal agencies, initiated COVID-19 mitigation activities in March 2020. Activities included educational sessions, training for infection prevention and control, health and safety assessments, and the development of standard operating procedures (SOPs). The Health Unit and US CDC India office initiated COVID-19 case investigations and conducted contact tracing. Between March 2020-July 2021, 636 COVID-19 cases (72% males), including 48 clusters (size range 2-10 cases), were reported. Overall case fatality rate was 1.5%. Of case patients, 82% (523) were Indians, and 18% (113) were Americans. On presentation, 22% (138/625) of cases were asymptomatic. The median time from symptom onset to notification to the Health Unit was three days (Interquartile range 1-5). The Health Unit identified 2,484 contacts (positivity rate 25%). Frequency of case presentation in the US Mission India closely resembled the pattern of COVID-19 waves in India. The attack rates ranged over the time period between 10-19%, the highest at 19% in Delhi.

**Conclusions:** COVID-19 mitigation strategies were implemented in collaboration with multiple agencies and helped prevent the transmission of COVID-19 and large COVID-19 clusters in the US Mission India.

## Introduction

Coronaviridae are an RNA family of viruses that have the tendency to jump from animals to humans. ^1^ In the past two decades, three human coronaviruses have emerged leading to acute respiratory illness and significant morbidity and mortality: severe acute respiratory syndrome coronavirus (SARS-CoV-1), Middle Eastern respiratory syndrome coronavirus (MERS-CoV), and most recently, COVID-19 coronavirus (SARS-CoV-2). In December 2019, SARS-CoV-2 first emerged in Wuhan, China. COVID-19, the disease caused by infection with SARS-CoV-2, was subsequently labeled as a pandemic by WHO in March 2020 due to its rapid rate of spread worldwide and associated mortality and complications^3^, including rare neurological delayed complications like transient ischemic attacks. ^4^

Between March 2020 and June 2021, over 30 million cases of COVID-19 were diagnosed in India, including two major waves in September-December 2020 and April-May 2021.^5^ During that period, over 600 COVID-19 cases occurred in the United States Diplomatic Mission to India (US Embassy and Consulates in India) American employees and their family members, Indian employees, contractors, and household staff. Throughout this period, as part of its COVID-19 public health response program, US Mission India implemented COVID-19 mitigation measures and case investigation with contact tracing for all cases. It further offered and administered COVID-19 vaccination to employees upon availability beginning in April 2021.

As part of an after-action-review exercise to inform future response efforts, we assessed the COVID-19 response across the five US Mission India posts (New Delhi, Chennai, Kolkata, Mumbai, and Hyderabad) to fill the gap in knowledge about COVID-19 transmission and mitigation in the unique community of US Mission sites in India. In this report, we describe the completeness, timeliness, and effectiveness of COVID-19 mitigation activities. We describe the COVID-19 cases in US Mission, including a comparison to cases in India overall and in specific consular cities between March 2020 and July 2021.

## Methods

### Study design

We drew the data from information collected through non-systematic COVID- 19 contact tracing and described the COVID-19 mitigation activities undertaken by US Mission India in 2020 and 2021. This was a secondary analysis of case investigation and contact tracing data collected by the Health Unit from March 2020 – July 2021.

### Study site

US Mission India facilities: Embassy New Delhi, Consulate General Mumbai, Consulate General Chennai, Consulate General Hyderabad, and Consulate General Kolkata.

### COVID-19 mitigation activities

We collected and compiled information about the US Mission’s COVID-19 response and mitigation activities during the study period. This included a review of policy and guidance documents and standard operating procedures; execution of trainings, webinars, COVID-19 group meetings, seminars, town halls, and vaccination drives; and development and dissemination of newsletter articles and management notices.

### Analysis of health unit case and contact investigation program

We included eligible participants from Delhi, Mumbai, Hyderabad, Kolkata, and Chennai sites.

#### Inclusion criteria

All US direct hires (USDH), eligible family members (EFM), locally employed staff (LES), local guard force (LGF), and others (housekeepers, nannies, and drivers working for Americans and contractors) who either tested positive for COVID-19 by rapid antigen test or Reverse Transcriptase Polymerase Chain Reaction (RT-PCR) / had a clinical diagnosis of COVID-19 while posted or during travel within India, or tested positive/ had a clinical diagnosis of COVID-19 while on a temporary personal or official visit to India during the study period.

#### Exclusion criteria

All USDH, EFM, LES, LGF, and others (housekeepers, nannies, and drivers working for Americans and contractors) who tested positive for COVID-19/ had a clinical diagnosis of COVID-19 while they were traveling outside of India for official or personal reasons.

A COVID-19 cluster was defined as two or more epidemiologically linked cases during the study period.

We abstracted the COVID-19 case data for US Mission cities in India (Delhi, Mumbai, Hyderabad, Kolkata, and Chennai Missions) from the Ministry of Health and Family Welfare (Government of India) website for comparison with reported US Mission cases in those respective cities.

### Data analysis

We calculated the distribution frequency of selected variables, including nationality, for each of the US Mission in India. Case investigation and contact tracing data included demographic data for both cases and contacts, date of symptom onset, date of contact by the Health Unit case investigation and contact tracing program, number of contacts, duration of isolation and outcome for cases, contact type (i.e., symptomatic or asymptomatic), and test results for contacts. For all COVID-19 cases, descriptive analysis was performed to assess age, sex, frequency of case identification by Mission (epidemic curve), contacts per case, positivity rate among contacts by type of contact (i.e., household, office, or social), and comparison of US Mission case trends with overall case trends in India and Consular cities. We used the Chi-square test to assess the association between selected categorical variables and calculated Odds Ratio (OR) with a 95% Confidence Interval (C.I). The data were analyzed using Epi Info Software, version 7.2.4.1 (US CDC).

### Ethical considerations

We have obtained approvals from US CDC (Atlanta) and Bureau of Medical Services (Washington, D.C.), US Department of State, for the secondary data analysis of pre-existing dataset of COVID-19 public health response at US Mission India. This study did not involve any primary data collection; hence consent from participants or parents in case of dependent children was not required. This review was deemed non-research by the Office of the Associate Director for Science of US CDC, hence a full Institutional Review Board approval not recommended. The dataset was obtained after stripping of all personal identifying information in order to maintain the confidentiality of the participants.

## Results

US Mission in India reported its first COVID-19 case in March 2020 from New Delhi. Between March 2020 and July 2021, a total of 636 COVID-19 cases (72% males), including 48 clusters, were reported from the US Mission – India (Figure 1). The median age was 42 years, with an interquartile range of 35-49 years. Overall, 82% (523) were Indians, and 18% (113) were Americans.

**Figure 1.**
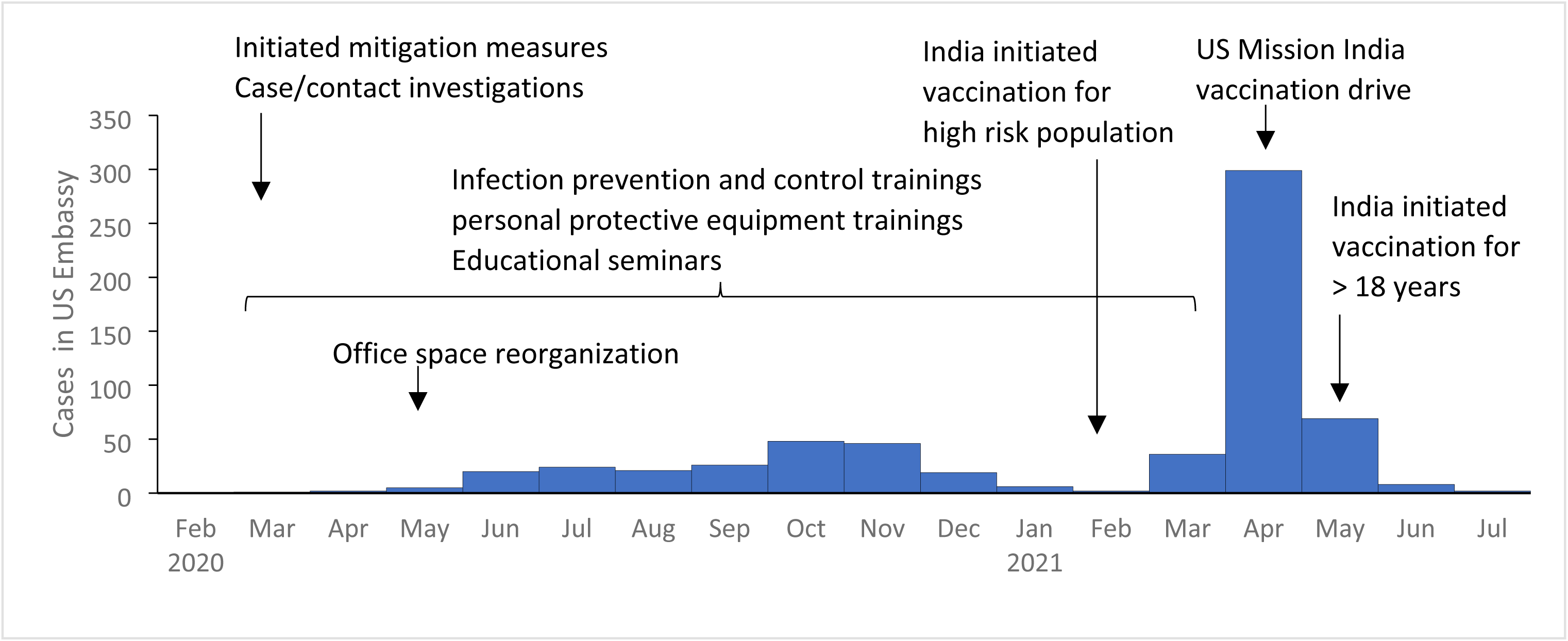
COVID-19 cases and mitigation activities in US Mission, India, Mar 2020-July 2021 (n=634)

### COVID-19 mitigation activities undertaken at US Mission in India

In response to COVID-19, the US Mission in India, in collaboration with internal US Mission divisions, initiated multiple activities, including education sessions for mission staff, hands-on training and demonstration for infection prevention and control, health and safety assessments, and issuing guidelines at regular intervals. In addition, US Mission Health Unit and US Centers for Disease Control and Prevention (CDC) India office initiated a program for COVID-19 case investigation and contact tracing follow-ups. (Figure 1)

The education sessions included health education and risk communication through open forum webinars and virtual town halls, which were conducted in both English and Hindi for better outreach. Periodic management notices and newsletter articles were shared with the Mission community, informing them about the sessions. Targeted infection prevention and control (IPC) training sessions included hands-on demonstrations for various sections of the workforce, including supervisors, contract cleaning staff, locally employed staff (LES), motor pool, and cafeteria staff. Other trainings included personal protective equipment training, cleaning and infection training, social distancing training, and hand washing training. Health and safety assessments were done for congregate workplace settings, and recommendations were provided for modified layouts to promote physical distancing, improved ventilation, and air circulation, and cleaning and sterilization guidance.

US Mission India Health Unit, CDC, and other agencies collaborated and developed guidelines for COVID-19 to maintain the continuity of the Mission’s critical activities. Workplace risk reduction guidelines included guidance on mask use, physical distancing, staying at home when ill, increased telework and virtual meetings, and temperature screening and symptom checking. Other guidance topics included monitoring and evaluating COVID-19 cases and contacts, COVID-19 case cluster identification and response, travel-associated guidance for domestic and international travel, and case isolation and contact quarantine support. Once the COVID-19 vaccine became available in April 2021, the Health Unit planned and implemented vaccination drives for all Mission staff and community.

Between May 2020 and July 2021, the US Mission collaborated with the COVID-19 working group experts to conduct a weekly “Doctors are In,” Webex seminar to address question from the US Mission community. To reduce the risk of COVID transmission in the workplace, the US Embassy Health Unit implemented the strategy of symptom self-screening. All employees were instructed to stay at home and update the Health Unit if they are sick, if anyone in their family (living in the same household) is sick, if they test positive for COVID-19, or if anyone in their family tests positive for COVID-19. Employees required clearance from a medical provider to return back to work. Numerous other mitigation measures were added, including temperature screenings, a mask mandate, installation of hand sanitizers (with foot-operated dispensers) outside every building, and the promotion of telework.

The core contact tracing team closely followed COVID-19 guidance in India and around the world to implement Indian guidance (the law of the land) in coordination with US guidance (being a US Mission). Where there was divergence, the US Mission followed the stricter of the two to ensure that both Indian and US guidance was followed. Familiarity with guidance in other countries was also important within this diplomatic community as some essential official travel took place even during the lockdowns.

We ensured that all cases and contacts completed government-recommended isolation or quarantine as appropriate, and underwent the recommended COVID-19 testing based on exposure and symptoms onset as per the guidelines.

### Descriptive analysis of the COVID-19 cases in US Mission in India

Of the reported 636 cases among the US Mission, the case fatality rate was 1.5% (10/636). Of the 10 deaths, eight occurred in 2021, nine were Indian, one was from the Chennai consulate, and nine were from the New Delhi Embassy.

The distribution of COVID-19 cases in the US Mission India closely resembled the pattern of COVID-19 cases in India. In 2020, the cases peaked during the first wave from October and November; similarly, the cases peaked in 2021 during the second wave from April-May 2021. COVID-19 case trends from the individual consulates mirrored their respective host city trends (Figure 2). The peak in cases in 2020 came early in Chennai, Hyderabad, and Mumbai from June-August compared to October-November in Delhi and Kolkata. In 2021, cases peaked in Delhi, Chennai, Hyderabad, Kolkata, and Mumbai in April. (Figure 2)

**Figure 2.**
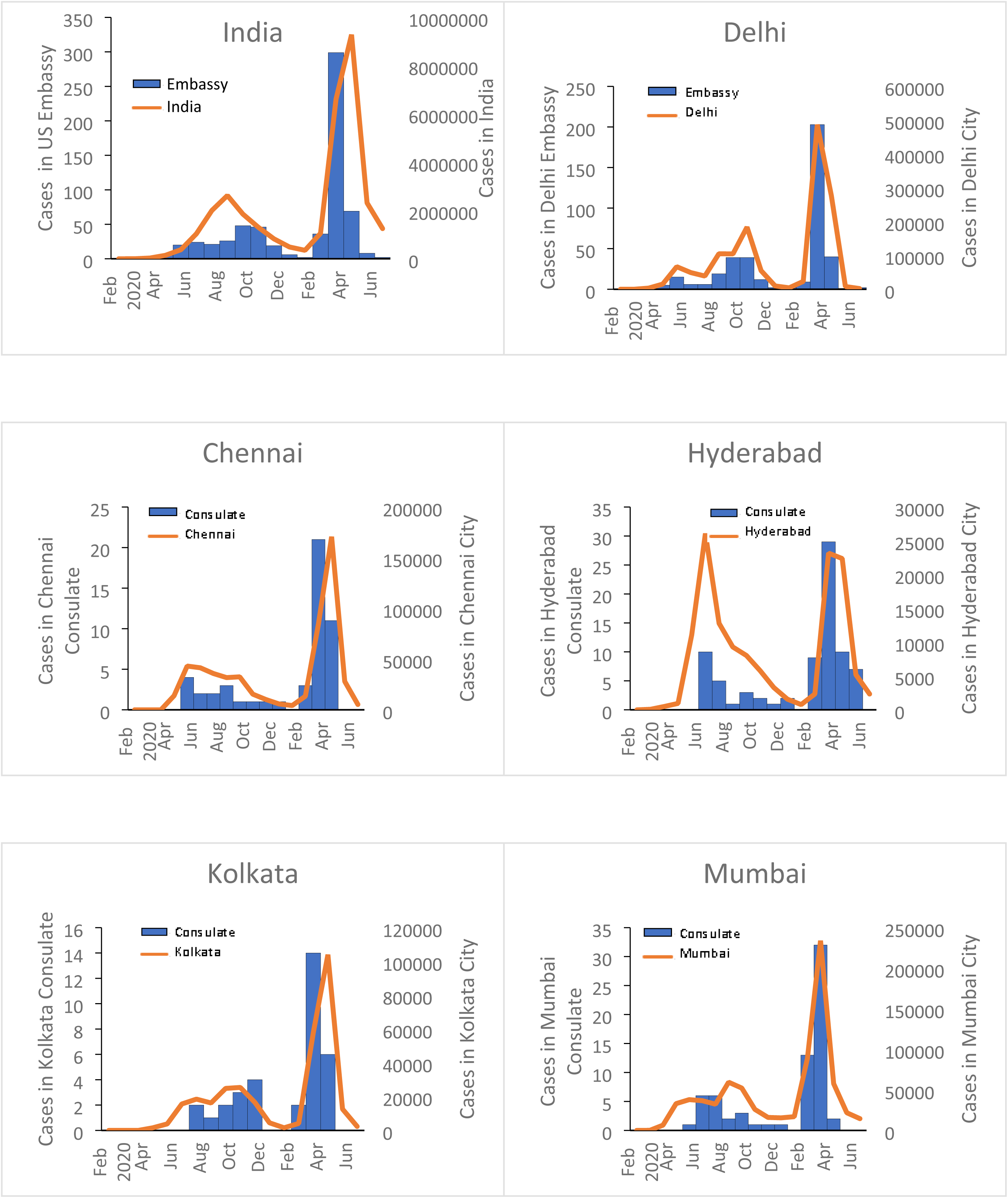
COVID-19 cases by month in US Mission, India, Mar 2020-July 2021 (n=634)

We identified and responded to 48 COVID-19 clusters across the US Mission in India. The median size of the cluster was 2 cases (range 2-10 cases), and 80% (38) clusters included ≤3 cases.

Comparing the COVID-19 cases among the consulates in 2020, we observed that attack rates among consulates ranged between 3-6%, the highest at 6% in Delhi. The overall attack rate did not differ between Americans (3%) and Indians (5%). However, in the Kolkata consulate, Americans had the highest attack rate of 16% compared to 4% among Indians. (Table 1)

In 2021, the attack rates among consulates ranged between 7-14%, the highest at 14% in Hyderabad. The overall attack rate is similar between Americans (10%) and Indians (10%). However, at the Kolkata consulate, Americans had the highest rate of 21% compared to 7% among Indians. Comparing the overall COVID-19 cases amongst consulates between June 2020- July 2021, attack rates ranged between 10-19%, the highest at 19% in Delhi. In the Kolkata consulate, Americans had the highest overall rate of 37% compared to 12% among Indians. However, the overall attack rate across all consulates was similar at 13% among Americans compared to 15% among Indians. (Table 1)

**Table 1.** COVID-19 Cases and attack rate in US Mission India, Mar 2020-July 2021 (n=636)

| Consulate | A.R. |  |  | A.R. |  |  | Total | N | AR % |
| --- | --- | --- | --- | --- | --- | --- | --- | --- | --- |
|  | American | n | % | Indian | n | % |  |  |  |
| Chennai | 2 | 92 | 2 | 12 | 405 | 3 | 14 | 495 | 3 |
| Delhi | 19 | 536 | 4 | 125 | 1984 | 6 | 144 | 2520 | 6 |
| Hyderabad | 1 | 86 | 1 | 21 | 320 | 7 | 22 | 406 | 5 |
| Kolkata | 3 | 19 | 16 | 11 | 249 | 4 | 14 | 268 | 5 |
| Mumbai | 3 | 145 | 2 | 17 | 563 | 3 | 20 | 708 | 3 |
| Total | 28 | 878 | 3 | 186 | 3521 | 5 | 214 | 4397 | 5 |

Year 2021
| Consulate | A.R. |  |  | A.R. |  |  | Total | N | AR % |
| --- | --- | --- | --- | --- | --- | --- | --- | --- | --- |
|  | American | n | % | Indian | n | % |  |  |  |
| Chennai | 7 | 92 | 8 | 29 | 405 | 7 | 36 | 495 | 7 |
| Delhi | 49 | 536 | 9 | 210 | 1984 | 11 | 259 | 2520 | 10 |
| Hyderabad | 8 | 86 | 9 | 49 | 320 | 15 | 57 | 406 | 14 |
| Kolkata | 4 | 19 | 21 | 18 | 249 | 7 | 22 | 268 | 8 |
| Mumbai | 17 | 145 | 12 | 31 | 563 | 6 | 48 | 708 | 7 |
| Total | 85 | 878 | 10 | 337 | 3521 | 10 | 422 | 4397 | 10 |

Year 2020-21
| Consulate | A.R. |  |  | A.R. |  |  | Total | N | AR % |
| --- | --- | --- | --- | --- | --- | --- | --- | --- | --- |
|  | American | n | % | Indian | n | % |  |  |  |
| Chennai | 9 | 92 | 10 | 41 | 405 | 10 | 50 | 495 | 10 |
| Delhi | 68 | 536 | 13 | 335 | 1984 | 17 | 403 | 2520 | 16 |
| Hyderabad | 9 | 86 | 10 | 70 | 320 | 22 | 79 | 406 | 19 |
| Kolkata | 7 | 19 | 37 | 29 | 249 | 12 | 36 | 268 | 13 |
| Mumbai | 20 | 145 | 14 | 48 | 563 | 9 | 68 | 708 | 10 |
| Grand Total | 113 | 878 | 13 | 523 | 3521 | 15 | 636 | 4397 | 14 |

People between the ages of 41-51 years had the highest caseload, while people above the age of 60 years had the highest case-fatality rate. (Table 2) At the time of presentation at the Med Unit, about 22% (138/625) of cases were asymptomatic. Among the 498 symptomatic cases, 63% reported fever, followed by body aches (48%), cough (43%), headache (30%), and sore throat (30%). (Figure 3)

**Figure 3.**
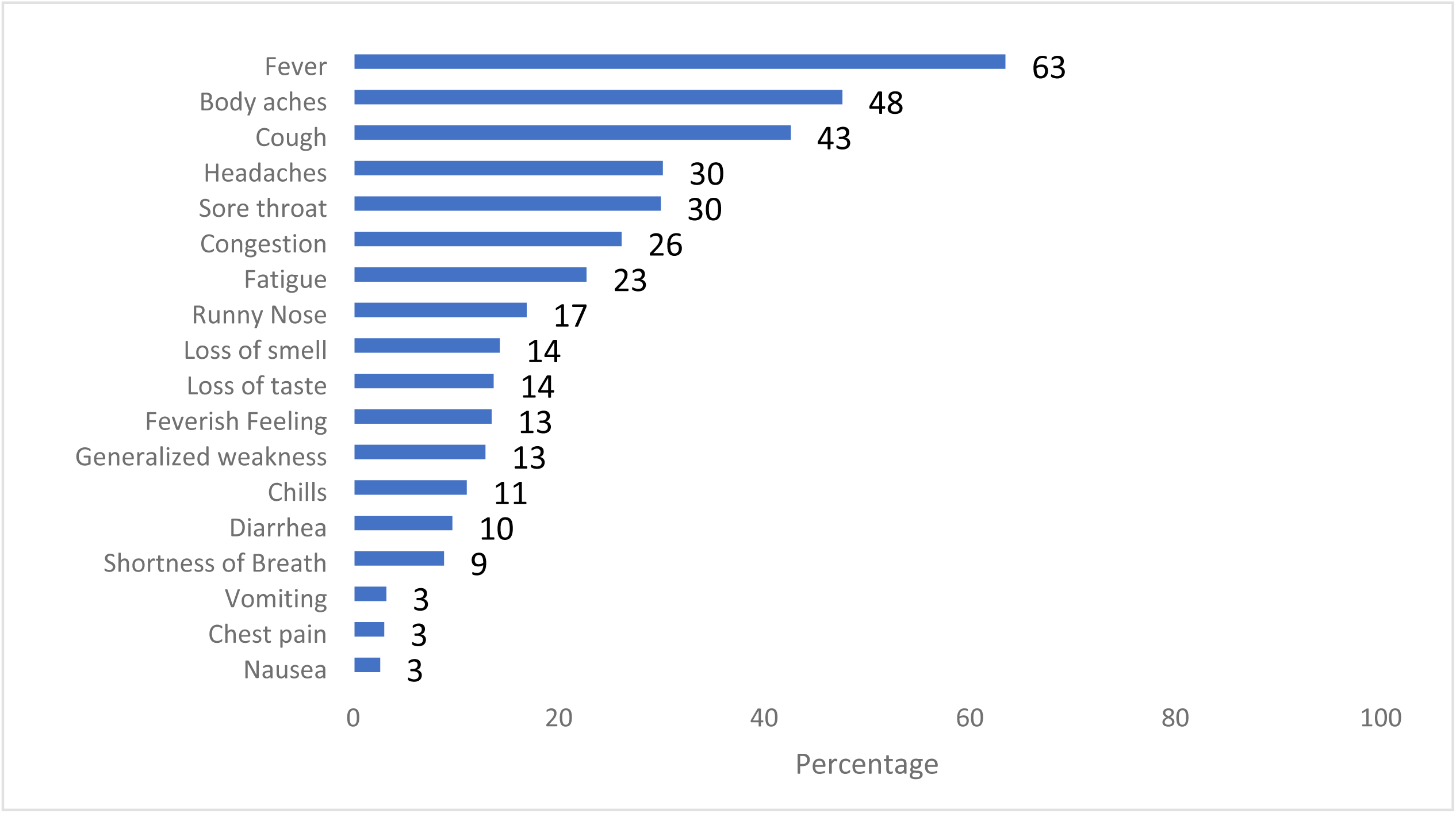
Symptoms at presentation among COVID-19 cases, US Mission India, March 2020 - July 2021 (n=498)

**Table 2.**
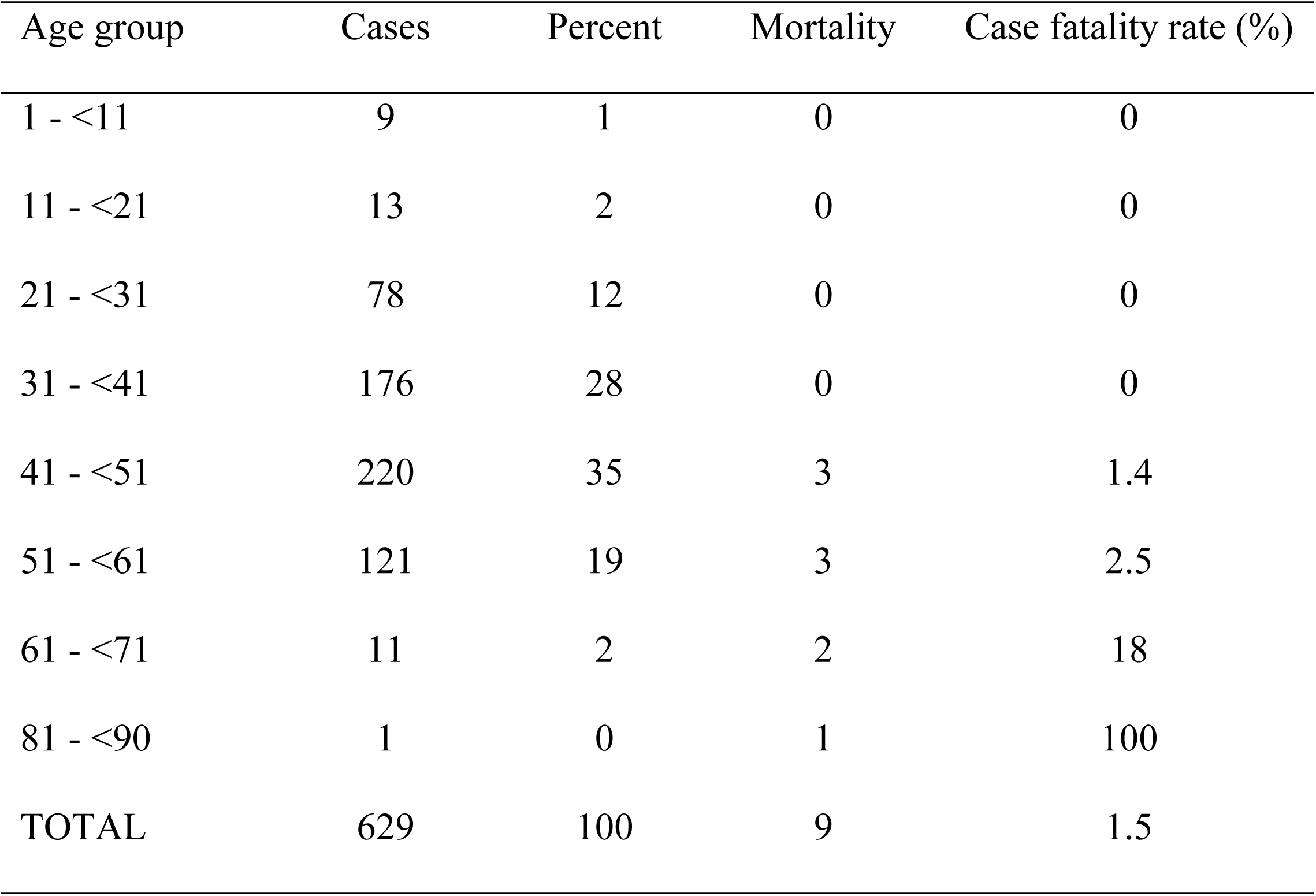
Age group wise cases and mortality rate among COVID-19 cases, US Mission India, March 2020 - July 2021.

Among the 627 cases who reported their smoking status, 15% (96/627) were smokers, 5% (34/627) were past smokers, and 10% (62/627) were current smokers. Overall, 27% (167/627) had comorbidities. Among 167 cases with comorbidities, 57% (96/167) reported hypertension, with a 5% (5/96) mortality rate (Table 3). Moreover, 2% (11/627) reported reinfection with COVID-19.

**Table 3.** Comorbidity among COVID-19 cases, US Mission India, March 2020 - July 2021 (n=167)

| Comorbidity | Number | % | Deaths | % |
| --- | --- | --- | --- | --- |
| Hypertension | 96 | 57 | 5 | 5 |
| Diabetes | 69 | 41 | 4 | 6 |
| Asthma | 50 | 30 | 1 | 2 |
| Heart Disease | 11 | 7 | 1 | 9 |
| Lung disease | 3 | 2 | 0 | 0 |
| Liver disease | 1 | 1 | 0 | 0 |
| Hypertension and diabetes | 37 | 22 | 4 | 11 |
| Any one comorbidity with age >45 years | 99 | 59 | 6 | 6 |
| Any two comorbidities with age >45 years | 38 | 23 | 5 | 13 |
| Any three comorbidities with age >45 years | 8 | 5 | 1 | 13 |
| Hypertension and diabetes with age >45 years | 25 | 15 | 4 | 16 |
| Hypertension, diabetes, and heart disease with age >45 years | 2 | 1 | 1 | 50 |

The case fatality rate (CFR) was 3.5% (8/230) among those age >45 years compared to 0.5% (2/398) among those < 45 years (Odds Ratio [OR] 7.1, CI:1.5-34, p=0.004). Among cases with both hypertension and diabetes, the CFR was 11% (4/37) compared with 1% (6/391) among those without either comorbidity (OR: 11.9, CI:3.2-44.5, p <0.0005). The fatality rate reported in US Mission India was 2.5 per 1000 population at risk (Tables 1 and 2). (New Delhi was 3.57 per 1000 population at risk, and Chennai was 2.01 per 1000 population at risk).

The median time from symptom onset to the notification (in days) to Health Unit in Mission India was three days (Interquartile range 1-5). The median time from symptom onset to COVID- 19 testing (n=491) was three days (interquartile range 2-15 days). The most frequently reported reason for the delay in notification to Health Unit in Mission India among cases was the perception of symptoms as fatigue (25%), delay from the local treating physician in suspecting COVID-19 (21%), and perception of initial sign to be a seasonal allergy (17%) or COVID-19 vaccine side effect (15%). (Figure 4)

**Figure 4.**
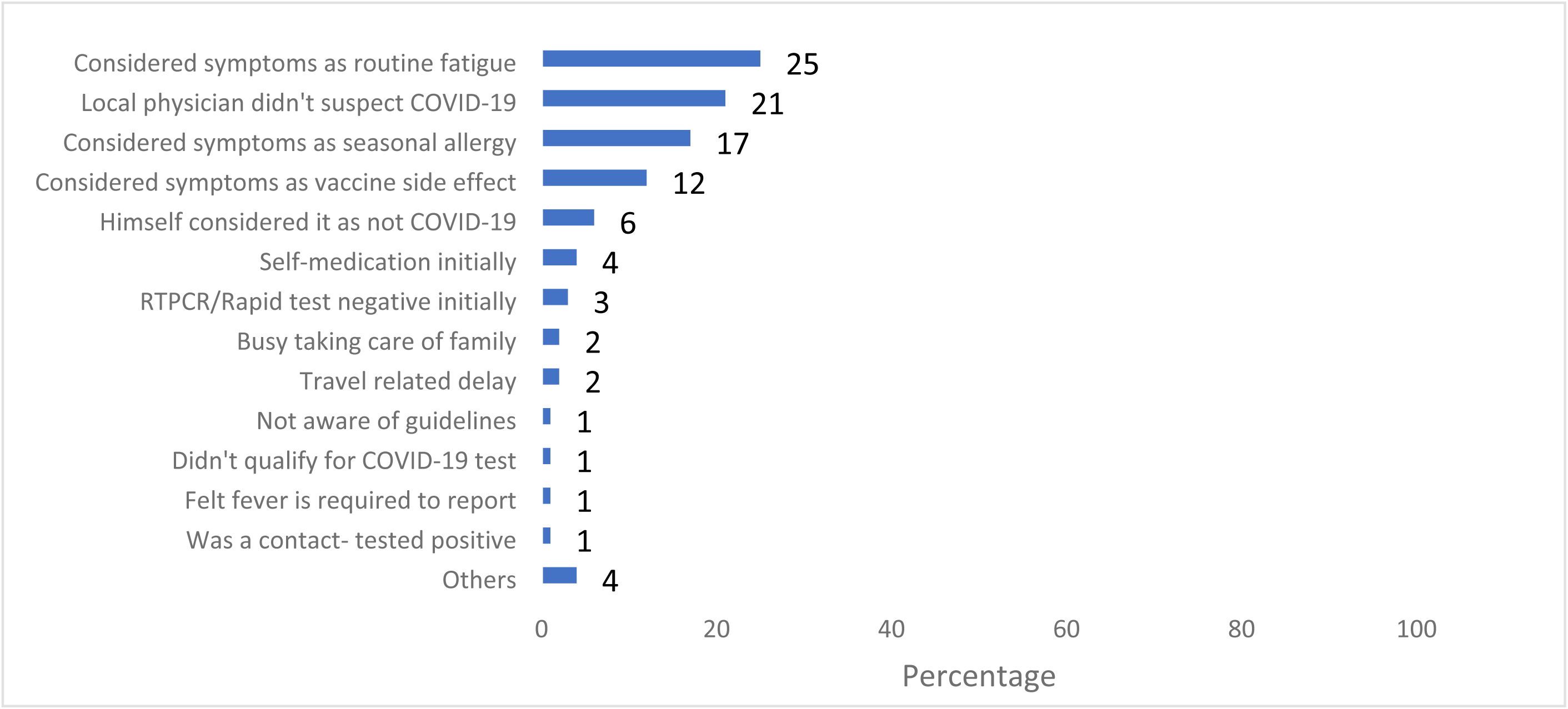
Perception among COVID-19 cases for delay in notification COVID-19 cases, at US Mission India, March 2020 - July 2021 (n=353)

Laboratory confirmation of the COVID-19 cases was mostly from RT-PCR (485/489), followed by a rapid antigen test (140/144) at a hospital or a private laboratory. Cycle Threshold Scores (C.T. score) for RT-PCR were available for 273 cases with RT-PCR (median 19 with an interquartile range of 17-21). Eleven cases were diagnosed clinically, and (47%) 21/45 had a chest x-ray for COVID-19. Overall, 8% (51/619) reported travel history within two weeks before the infection; 44 Domestic travel (14 in 2020 and 30 in 2021) and 11 international travel (1 in 2020 and 10 in 2021).

Two third of the cases received Vitamin C, Zinc, and Vitamin D as part of the COVID-19 treatment, and half received Ivermectin and Azithromycin. (Figure 5) Hospitalization was 7% (45/627); 44 Indian and 1 American. 6% (20/326) who received Ivermectin as part of COVID-19 treatment got hospitalized compared to 8% (25/310) who did not receive Ivermectin showing no statistically significant difference (Odds Ratio 0.7, C.I:0.4-1.37, p 0.8). Indians were following the Government of India’s Treatment guidance while Americans followed US guidelines.

**Figure 5.**
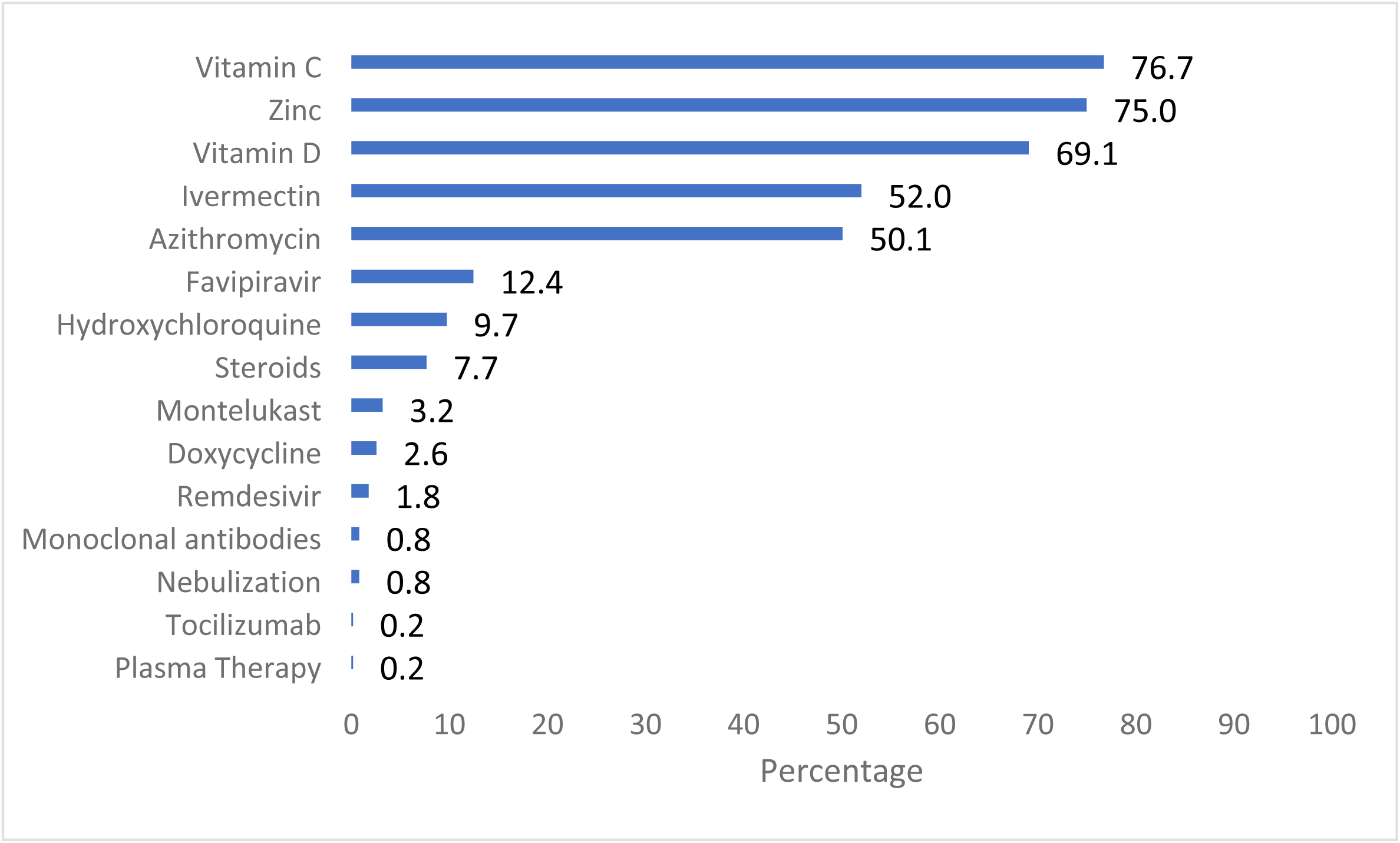
Treatment history among COVID-19 cases US Mission India, March 2020 - July 2021 (n=627)

All cases were isolated (14 days initially) as per the Ministry of Health and Family Welfare, Government of India guidelines, and contact tracing was done for each reported case. All close contacts were recommended quarantine for 14 days (per Government of India policy), monitored for COVID-19 symptoms, and tested as per the Government of India recommended guidelines on the fifth day following exposure. Of the 636 reported cases, the Health Unit conducted case investigations and found 2,484 contacts between March 2020-July 2021. The overall contact positivity rate was 25% (range 20-26% among all consulates). Office contact positivity rate was highest in Hyderabad (14%), household contact positivity rate highest in Kolkata (70%), social contact positivity rate highest in Delhi (9%), and family contact positivity rate was highest in Mumbai 43%. (Table 4)

**Table 4.** Contact positivity among COVID-19 cases US Mission India, March 2020 - July 2021 (n=627)

| Consulates | Office |  |  | Household |  |  | Social |  |  | Family |  |  | Total |  |  |
| --- | --- | --- | --- | --- | --- | --- | --- | --- | --- | --- | --- | --- | --- | --- | --- |
|  | n | +ve | % | n | +ve | % | n | +ve | % | n | +ve | % | n | +ve | % |
| Chennai | 29 | 1 | 3 | 4 | 0 | 0 | 0 | 0 | 0 | 100 | 33 | 33 | 133 | 34 | 26 |
| Delhi | 390 | 37 | 9 | 103 | 15 | 15 | 56 | 5 | 9 | 1173 | 394 | 34 | 1722 | 451 | 26 |
| Hyderabad | 136 | 19 | 14 | 11 | 0 | 0 | 13 | 0 | 0 | 169 | 48 | 28 | 329 | 67 | 20 |
| Kolkata | 23 | 1 | 4 | 10 | 7 | 70 | 1 | 0 | 0 | 88 | 24 | 27 | 122 | 32 | 26 |
| Mumbai | 91 | 2 | 2 | 11 | 5 | 45 | 0 | 0 | 0 | 76 | 33 | 43 | 178 | 40 | 22 |
| Overall | 669 | 60 | 9 | 139 | 27 | 19 | 70 | 5 | 7 | 1606 | 532 | 532 | 2484 | 624 | 25 |

Among the 530 who reported their vaccine history till July 2021, 74% had taken Moderna, 10% Covishield and the rest had other vaccines (Covaxin and Sputnik) available in India. Vaccination started in India in January 2021 India (initially, only high-risk individuals or those above 60 years were eligible for vaccination). US Mission in India initiated vaccination for all staff in April 2021.

## Discussion

COVID-19 mitigation strategies were well planned and implemented in collaboration with multiple agencies in the US Mission India. Although the case investigation and contact tracing program led by the Health Unit New Delhi was labor intensive, it went a long way in preventing the transmission of COVID-19 and large COVID-19 clusters in the US Mission India. Cases peaked during the waves in each city, which was as expected, but the Mission numbers stayed low.

One-fifth (20%) of the cases at presentation were asymptomatic in our analysis; in previous analyses, this percentage was found to be anywhere between 13% and 31%. ^6,7,8^ One-third of cases had comorbidities. Case fatality remained low in the US Mission in comparison with overall rates in each respective city and India’s national average. US Mission India’s COVID-19 mitigation strategies and the effectiveness of the contact tracing program likely played a role in low case fatality rates.

Most deaths reported within the US Mission occurred in the US Embassy New Delhi, but that can be attributed to the large size of the US Embassy in New Delhi compared to the other Consulates in India. The US Mission countrywide and state-wide mortality ratios were lower compared to those for India and the Consulate cities^9^, as well as compared with estimates published elsewhere^10 11^. These findings were observed despite the fact that the study population included mostly US Mission India staff, who were working and travelling domestically and internationally throughout the majority of the study period.

Americans at the Kolkata consulate had a higher risk of contracting COVID-19 compared with Americans at other US mission cities both overall and in respective years of analysis, with double the risk in 2021. One possible explanation for this could be that the small size of the American community in Kolkata compared with the US Embassy in New Delhi (19 vs. 536) offered less opportunity for social distancing.

Our analysis shows there are 11 times more chances of mortality amongst individuals aged >45 with co-morbidities Hypertension and Diabetes. The presence of chronic conditions such as coronary heart disease, diabetes, and hypertension increases the risk of severe complications of COVID-19 disease ^12,13,14,15,16^, with a finding also seen in our analysis. Another point worth noting is the challenges faced at the peak of second COVID-19 wave in New Delhi, including bed crisis and oxygen crisis.^17^

Our analysis found that one of four contacts became COVID-19 positive. Martinez-Fierro et al. ^18^ found that 42% of contacts tested positive in Mexico. Given the high positivity rate among contacts, early detection. and isolation is the key, as often the contacts can be asymptomatic but are transmitting COVID-19 to others.^19^

Reasons for delayed notification to the Health Unit suggested that individuals whose occupations required physical labor, such as facility workers (electricians, plumbers, construction workers, masons, carpenters, welders) and security guards, often felt they were overworked and ignored fatigue and body aches. Also, physicians often waited for fever to appear before recommending a COVID-19 test, even though the Government of India (Ministry of Health and Family Welfare) did not have fever as an essential criterion for diagnosing COVID-19. Even in our analysis, fever was only present in 63% of the COVID-19 positive patients. The existing literature supports that fever alone is not a specific indicator of COVID-19 infection, with Schneider et al.^20^ suggesting that body temperature may be an insufficient indicator of COVID-19 infection and Leal et al. ^21^ offer fever as an essential component of a trio of symptoms (fever, anosmia, and ageusia) with high specificity for COVID-19 diagnosis.

These findings will be helpful to US Mission India and other US Missions globally, the Indian Ministry of Health, and other US and Indian government agencies and organizations involved in protecting and improving public health. The findings from this analysis will help implement and strengthen the existing COVID-19 mitigation activities for current and future pandemic threats in US Mission India. While COVID-19 clinical outcomes in diplomatic missions have been described elsewhere ^22^, our analysis describes COVID-19 trajectory, outcomes, and mitigation measures, which have not been analyzed amongst any other diplomatic missions. These results can be adapted and contextualized for use in other similar populations/organizations.

### Limitation

We performed a secondary analysis on an existing data set primarily collected for non-systematic COVID-19 contact tracing, with the primary purpose of informing clinical care. As such, some variables of interest were missing, including symptom duration and genomic investigation and analysis. This limited our ability to assess the full clinical spectrum and compare circulating variants within the US Mission community to those circulating in the Consular cities.

### Conclusion and recommendations

COVID-19 mitigation strategies in US Mission India were implemented in collaboration with multiple agencies and helped prevent the transmission of COVID-19 and large COVID-19 clusters in the US Mission India.

We recommended to prioritize vaccination amongst those >45 years and having multiple comorbidities (hypertension, diabetes, and heart disease). These specific populations are at higher risk for severe complications, including death, and may benefit from targeted mitigation strategies.

As pandemics change, mitigation guidelines need to be revisited to ensure that they are appropriate and enforceable. Individuals having occupational and physical demands like facility workers (electricians, plumbers, construction workers, masons, carpenters, welders) and security guards should have targeted education with lower identified thresholds for seeking medical care, and providers should have lower thresholds for suspecting COVID-19 in workers in these professions. Future analysis could include long-term follow-ups, which will be more informative in terms of the long-term effects of COVID-19.

During the Pandemic times, we demonstrated a collaborative effort among clinicians and public health experts to tackle COVID-19 by preventing the disease rather than just treating the disease. Subsequently, clinicians worked as and alongside public health specialists to achieve lower attack and case fatality rates than expected in a unique population. Lessons learned from this pandemic response can be applied to future pandemics.

## Data Availability

Data submitted with the manuscript

## Acknowledgment

We acknowledge and thank all the US Mission India Staff who contributed to COVID-19 control and mitigation response.

## Disclaimer

The views expressed in this article are authors’ own and not necessarily those of the U.S. Government or US Department of State, Bureau of Medical Services or US Centers for Disease Control and Prevention.

